# SARS-CoV-2 infection and transmission in educational settings: cross-sectional analysis of clusters and outbreaks in England

**DOI:** 10.1101/2020.08.21.20178574

**Authors:** Sharif A Ismail, Vanessa Saliba, Jamie Lopez Bernal, Mary E Ramsay, Shamez N Ladhani

## Abstract

**Background:** There are limited data on SARS-CoV-2 infection and transmission in educational settings. This information is critical for policy makers and practitioners to ensure the safety of staff, students and the wider community during the COVID-19 pandemic.

**Methods:** Public Health England initiated enhanced national surveillance following the reopening of educational settings during the summer mini-term on 01 June 2020. COVID-19 related situations in educational settings across England were reviewed daily and followed-up until 31 July 2020. SARS-CoV-2 infection and outbreak rates were calculated for staff and students attending early year settings, primary and secondary schools during June 2020.

**Findings:** There were 67 single confirmed cases, 4 co-primary cases and 30 COVID-19 outbreaks during June 2020, with a strong correlation between number of outbreaks and regional COVID-19 incidence (0.51 outbreaks for each SARS-CoV-2 infection per 100,000 in the community; p=0.001). Overall, SARS-CoV-2 infections and outbreaks were uncommon across all educational settings. Staff members had an increased risk of SARS-CoV-2 infections compared to students in any educational setting, and the majority of cases linked to outbreaks were in staff. The probable transmission direction for the 30 confirmed outbreaks was: staff-to-staff (n=15), staff-to-student (n=7), student-to-staff (n=6) and student-to-student (n=2).

**Interpretation:** SARS-CoV-2 infections and outbreaks were uncommon in educational settings during the first month after the easing of national lockdown in England. The strong correlation with regional SARS-CoV-2 incidence emphasises the importance of controlling community transmission to protect educational settings. Additional interventions should focus on reducing transmission in and among staff members.

**Funding:** none

## INTRODUCTION

The lack of vaccines or effective treatment against severe acute respiratory syndrome coronavirus 2 (SARS-CoV-2) have forced many countries to impose strict lockdown measures, including school closures, to reduce the spread of the virus. Educational settings that have remained open during the coronavirus disease 19 (COVID-19) pandemic have enforced significant constraints on activities.^1,2^ There are concerns, however, that school closures have adverse consequences on children’s educational needs and social and mental well-being.^2^ In addition, children from disadvantaged backgrounds are more likely to suffer from school closures for a number of reasons, including access to free school meals and social services.^3^ This is particularly important in the context of young children having a significantly lower incidence of COVID-19 compared to adults and developing mainly transient and mild illness, with severe disease and fatalities rarely occurring in this age-group.^1,4^

In England, the first imported cases of COVID-19 were identified in late January 2020 and started to increase rapidly from early March and before plateauing in mid-April and then declining gradually.^5^ As part of the lockdown, schools across England were closed from 20 March 2020 with wider lockdown measures announced on 23 March 2020.^6^ Children of key workers, however, could attend school throughout the pandemic. The gradual easing of lockdown measures in England began on 10 May and, from 01 June 2020, included the partial re-opening of preschools and primary schools with children attending nursery, reception, year 1 and year 6 allowed to return, following implementation of strict infection control measures.^7^ Re-opening of educational settings was, however, partial (nursery, reception, year 1 and year 6 in primary schools, and years 10 and 12 in secondary schools), not mandatory and the decision to re-open schools was met with mixed responses from educational staff and parents. Consequently, not all schools re-opened and not all parents sent eligible children to school during the remainder of summer term (June to mid-July 2020)

Public Health England (PHE) has been monitoring and managing outbreaks of COVID-19 across all settings throughout the pandemic in England. Here, we summarise the cases, clusters and outbreaks of COVID-19 in educational settings during the month of June, focussing particularly on the index case, the potential source of infection, additional possible and confirmed COVID-19 cases within the same educational setting and public health measures implemented to control the spread of the virus.

## METHODS

In England, PHE is responsible for surveillance and public health management of COVID-19 in the community, including educational settings. Institutions are required to inform their local PHE Health Protection Team (HPT) of suspected or confirmed cases of COVID-19. HPTs then perform a risk assessment for each event and decide on additional investigations and measures required. HPTs record all events on HPZone, an online national database used to record health protection-related events that require public health management.

### Definitions

Educational settings included nurseries, preschools, infant schools, junior schools, primary schools, secondary schools, further education colleges, and settings catering to children of mixed age groups, or those of any age with special educational needs and/or disabilities (SEND).^8^ A case of COVID-19 was confirmed if there was a verifiable, laboratory result for a positive SARS-CoV-2 RT-PCR from an upper respiratory tract sample. Co-primary cases were defined as ≥2 confirmed cases with a common epidemiological link who were diagnosed at the same time, typically asymptomatic siblings diagnosed as part of contract tracing after their parent tested positive for SARS-CoV-2. An outbreak was defined as ≥2 epidemiologically linked cases, where sequential cases were diagnosed within a 14-day period. Within the educational setting, a “bubble” was defined as a group of staff and pupils who stay together, perform all activities together and do not interact physically or socially with other bubbles.^7^ The aim was to isolate individual bubbles if any member of the bubble became unwell, whilst allowing the remaining bubbles to continue to attend the educational setting.

### Investigation of clusters and outbreaks

A national meeting was held each day to discuss situations of interest across England, including cases and outbreaks in educational settings. A risk assessment was performed and a decision for extensive testing was made on a case-by-case basis, with a particular focus on situations where wider transmission might have occurred.

### Data management and analysis

To monitor situations in educational settings, all potential events reported across England were extracted and updated every day from HPZone and entered into a pre-developed MS Excel template. All situations reported to HPTs in June 2020 were followed up for at least 14 days after the educational settings closed for the summer term. Missing information was obtained from outbreak reports or directly from the local HPT or educational setting as required.

Data are mainly descriptive. To calculate average, weekly regional prevalence of SARSCoV-2 infection, we utilised national daily reports of positive SARS-CoV-2 RT-PCR results collated by PHE for COVID-19 surveillance. SARS-CoV-2 positive tests performed as part of Pillar II testing in the community were used to estimate regional prevalence and correlated with regional COVID-19 outbreaks in educational settings during June 2020.^9^ Linear regression analysis was used to assess correlation between outbreaks in educational settings and regional population, population density and community COVID-19 incidence in England during June 2020. In addition, SARS-CoV-2 infection rates were calculated for staff and students attending an educational setting during June 2020, irrespective of whether the infection was acquired within or outside the educational setting. Coprimary cases were counted as separates cases. Educational settings and students were categorised into:^10^ early years (nurseries and preschools for < 5 year-olds), primary schools (reception and school years 1 and 6 for 5–11 year-olds), and secondary schools (school years 10 and 12 for 12–18 year-olds). Schools for mixed ages and SEND schools that include children of all ages were not included in the SARS-CoV-2 infection rate estimates. Denominators were obtained from Department for Education (DfE) online reports.^11^ Student denominators did not include a small proportion of children of key workers or vulnerable children who attended school but were not part of the year groups returning to school in June 2020. Staff attendance was only available for primary and secondary schools and did not distinguish between student-facing or other staff members.

## RESULTS

In England, educational settings re-opened on 01 June 2020, starting with nursery, reception, year 1 and year 6 pupils (**Supplement Figure S1**); and extending to years 10 and 12 in secondary schools, sixth form and further education colleges from 15 June 2020 – although attendance at these latter settings was much lower than in early years settings and primary schools (**Supplement Figure S2**). Educational settings implemented strong infection control measures, including smaller classes separated into distinct social bubbles, maintaining social distancing where possible, and frequent hand washing among other measures. From 01 to 30 June 2020, the number of open educational settings in England rose from 20,500 to 23,400 and the number of children attending any educational setting increased from 475,000 to 1,646,000 (**Supplement Table S3**).

During the same period, PHE received 170 reports of COVID-19 related events in educational settings in England. Public health investigations did not identify any SARS-CoV-2 infections in 69 (40%) of these reports. The remaining 101 (60%) events involved a single confirmed case (n=67, 39%) co-primary cases (n=4, 2%) and confirmed outbreaks (n=30, 18%). Across all 101 situations, there were a total of 70 confirmed cases in children, and 128 in members of staff. Single confirmed cases were most prevalent in the East of England and confirmed outbreaks in the Yorkshire and Humber region during this period (**Figure 1**). Outbreaks, where they occurred, were usually small in size and 53% of confirmed outbreaks involved only one secondary case linked to the index case. Where the index case was a child, the maximum number of secondary cases was 2 (compared with 9 for staff members). Larger outbreaks were predominantly in early years settings and primary schools, as would be expected considering these are the settings most children returned to as partial school reopening occurred – see **Figure 2**). There was a strong correlation (R^2^=0.82, p=0.001) between outbreaks in educational settings and regional COVID-19 incidence, with an increase of 0.51 outbreaks for each SARS-CoV-2 confirmation per 100,000 in the community (standard deviation, 0.07; p=0.001) (**Figure 3**). There is no association between single cases in educational settings and regional incidence (p=0.52), between outbreaks and regional population size (p=0.75), or between single cases in educational settings and regional population size (p=0.42).

**Figure 1:**
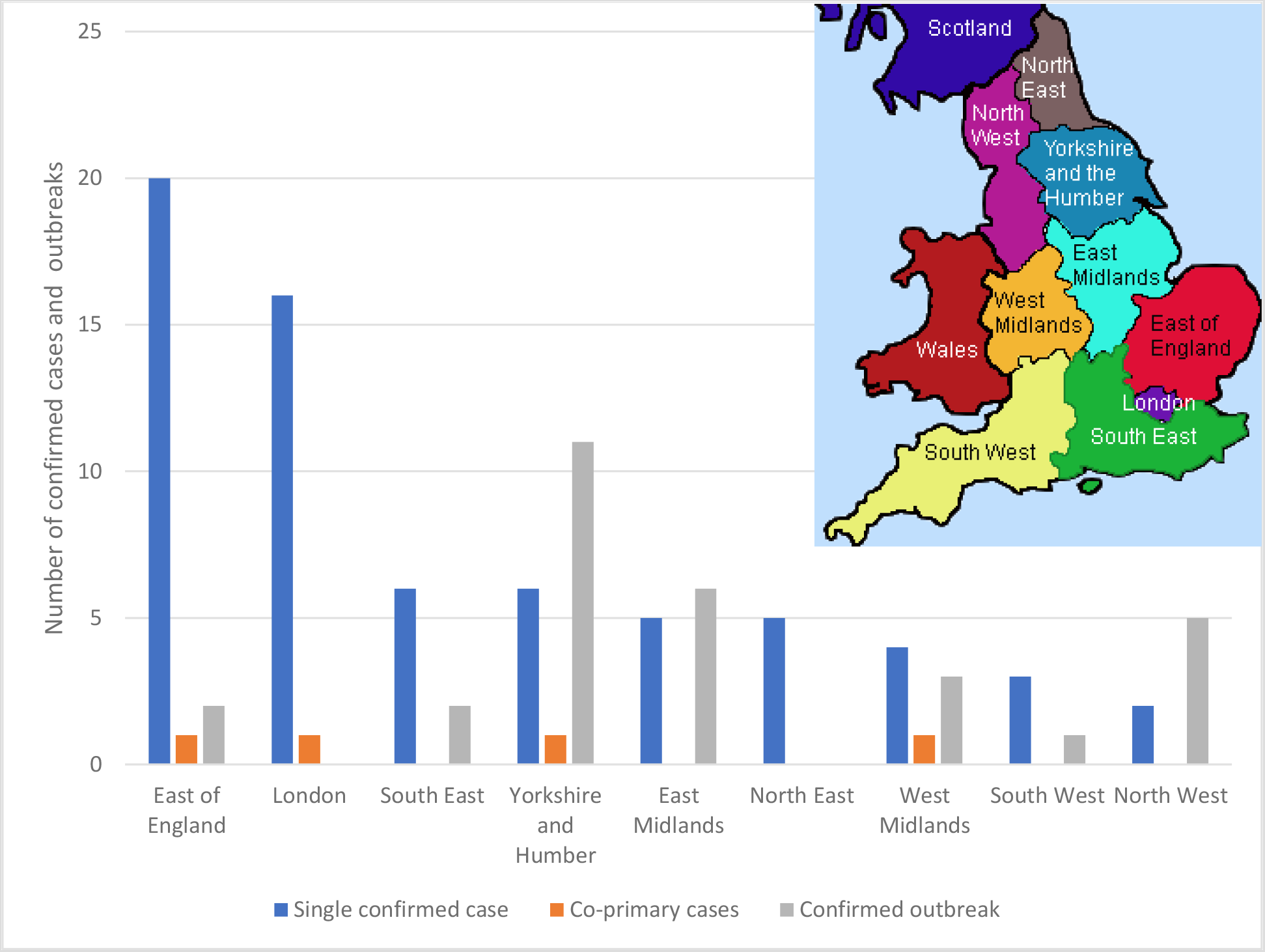
Regional distribution of single confirmed cases, co-primary cases (clusters) and confirmed outbreaks across England, June 2020. Figure for regions from: http://projectbritain.com/regions/index.htm

**Figure 2:**
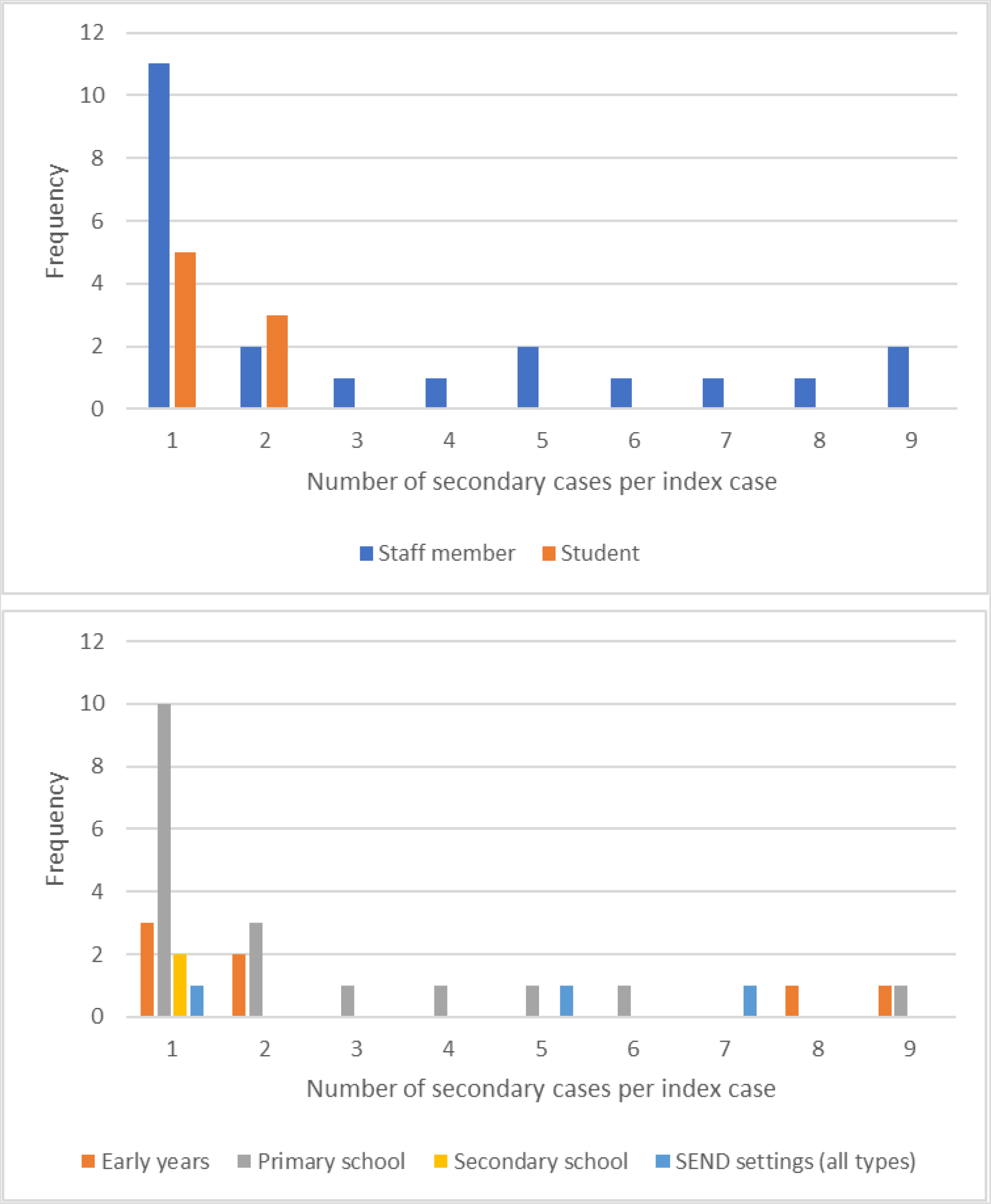
Number of secondary cases for confirmed outbreaks in educational settings in England in June 2020, by (i) index case (top), and (ii) type of educational setting (bottom)

**Figure 3.**
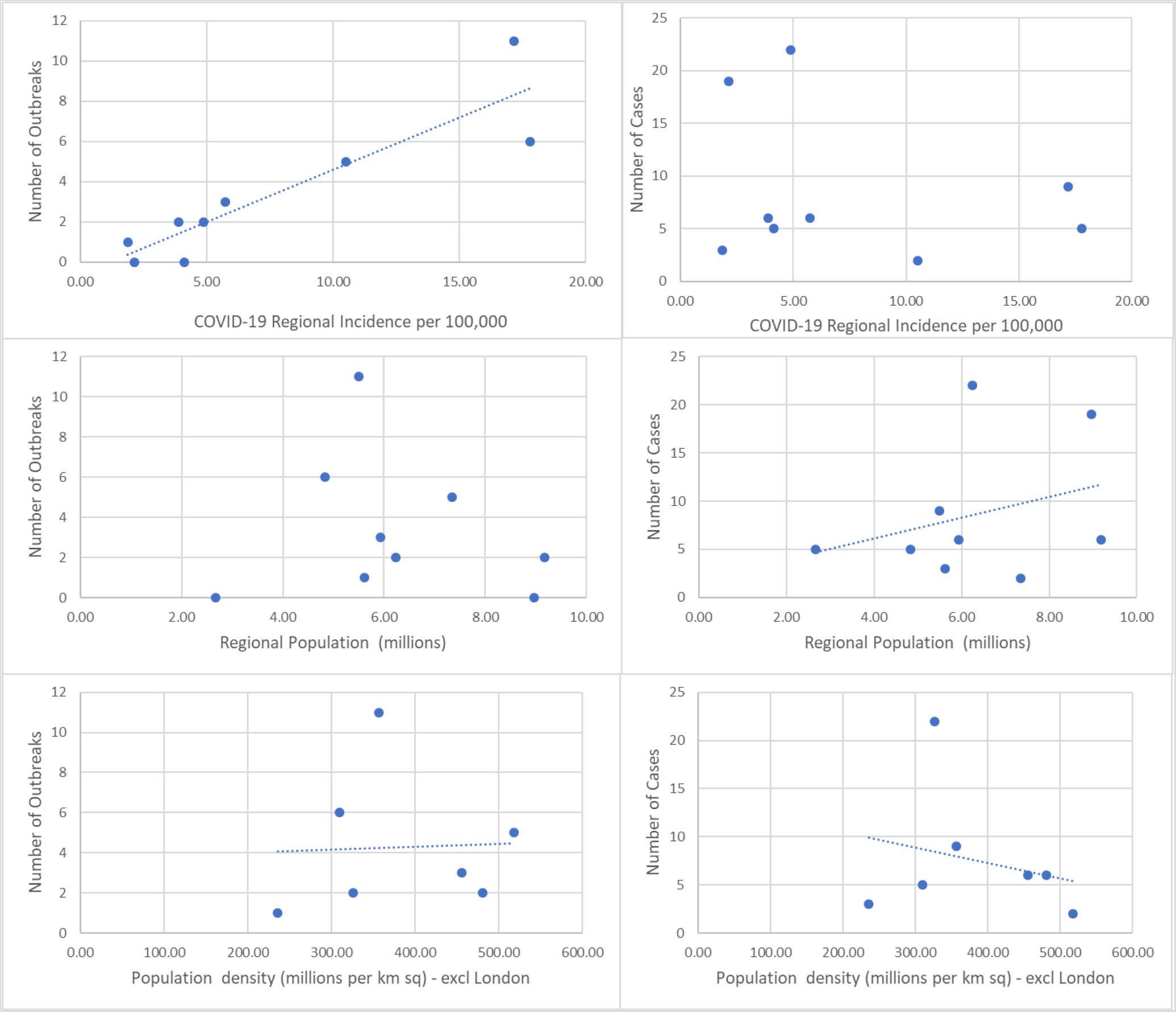
Correlation between number of outbreaks and regional SARS-CoV-2 incidence (per 100,000 population), regional population (millions) and population density (millions per km sq). *The population density graph excludes London which has a very high population density but no outbreaks*

### Risk of SARS-CoV-2 infection

The risk of SARS-CoV-2 infection and COVID-19 outbreaks was very low across all educational settings, with the highest risk in primary schools (4.8 events/1,000 schools/month), likely reflecting the distribution of educational settings that opened after the lockdown (**Table 1**). Staff had higher rates of individual SARS-CoV-2 infection and outbreaks than students, albeit with wide confidence intervals. Among students, the rate of individual infections was higher in primary school students but cases in children in early year settings were more likely to be linked to outbreaks.

**Table 1:** SARS-CoV-2 infection rates for cases, co-primary and outbreak events in educational settings, June 2020, England

|  | Number of Settings open |  |  | Numbers confirmed:<br>Staff/Student/Both |  |  | Rate per 1,000 settings per month |  |  |  |
| --- | --- | --- | --- | --- | --- | --- | --- | --- | --- | --- |
| Setting | Minimum | Maximum | average/d | Single | Coprimary | Outbreak | single | coprimary | outbreak | Total |
| Early years | 28,000 | 40,000 | 35,571 | 5/5 | 0/1 | 2/0/5 | **0.31**<br>(0.15-0.55) | **0.03**<br>(0.0007-0.16) | **0.20**<br>(0.007-0.20) | **0.51**<br>(0.05-0.80) |
| Primary | 6,900 | 18,100 | 13,845 | 24/21 | 0/3 | 9/2/7 | **3.5**<br>(2.6-4.6) | **0.22**<br>(0.04-0.63) | **1.3**<br>(0.77-2.1) | **4.8**<br>(0.20-1.04) |
| Secondary | 2,900 | 4,400 | 3,800 | 2/2 | 0/0 | 2/0/0 | **1.1**<br>(0.29-2.7) | **0** | **0.53**<br>(0.06-1.9) | **1.6**<br>(0.58-3.4) |
|  | Numbers attending |  |  | Student Numbers |  |  | Rate per 100,000 students per day |  |  |  |
| Settings | Minimum | Maximum | average/d | Single | Coprimary | Outbreak | Single | Coprimary | outbreak | total |
| Early years | 108,000 | 320,000 | 222,286 | 5 | 3 | 14 | **2.2**<br>(0.73-5.3) | **1.3**<br>(0.28-3.9) | **6.3**<br>(3.4-10.6) | **9.9**<br>(6.2-15.0) |
| Primary | 195,000 | 830,000 | 519,727 | 21 | 7 | 15 | **4.0**<br>(2.5-6.2) | **1.3**<br>(0.54-2.8) | **2.9**<br>(1.6-4.8) | **8.3**<br>(6.0-11.1) |
| Secondary | 78,000 | 126,000 | 101,417 | 2 | 0 | 0 | **2.0**<br>(0.24-7.1) | **0** | **0** | **2.0**<br>(0.24-7.1) |
|  | Numbers attending |  |  | Staff numbers |  |  | Rate per 100,000 staff per day |  |  |  |
| Staff * |  |  | 519,590 | 31 | 0 | 76 | **6.0**<br>(4.1-8.5) | **0** | **14.6**<br>(11.5-18.3) | **20.6**<br>(16.9-24.9) |

\* includes teachers (n=221,680) and non-teaching staff including teaching assistants (n=297,901)
\*includes coprimaries
staff

**Table 2:** Summary of wider testing for SARS-CoV-2 in selected educational settings, June 2020, England

| Region in England | Description of the situation | Children/ Staff attending | School closed? | Teachers and other staff |  |  | Students |  |  | Overall assessment |
| --- | --- | --- | --- | --- | --- | --- | --- | --- | --- | --- |
|  |  |  |  | Exposed | Positive | Pending/ unknown | Exposed | Positive | Pending/ unknown |  |
| Yorkshire and Humber | • 8 confirmed cases (7 staff, 1 child) in a special needs secondary school; only 1 staff member was symptomatic.<br>• Wider swabbing undertaken because of vulnerability of the population. | 16/24 | No | 22 | 6/18 | 0 | 16 | 1/12 | 0 | Confirmed outbreak mainly affecting staff in a special needs school, with evidence of transmission to a child. |
| East of England | • 13 confirmed cases (9 pupils, 4 staff) in an infant school<br>Staff member was the index case, 2 children then became symptomatic; infection source not known<br>• Partner of index case then tested positive.<br>• 4 bubbles in school, 2 affected. Bubble 1: 20 children, 3 staff; Bubble 2: 17 children. Only known point of contact between the two bubbles was shared pupil washroom facilities.<br>• Wider swabbing for all staff, students and household members: 165 tested; 9/41 children, 4/12 staff and 10/112 household members positive | 64/12 | Yes | 10 | 4/12 | 0 | 64 | 9/38 | 3 | Outbreak in an infant school, with staff member as index case. Evidence of wider transmission across 2 bubbles and within households. |
| Yorkshire and Humber | • 13 confirmed cases (3 pupils, 10 staff) in a primary school; 2 teachers (partners) were index cases. Infection source not known<br>• 2 main bubbles: Bubble 1: lunchtime supervision (14 pupils, 5 staff). Bubble 2: breakfast club (9 pupils, 1 staff)<br>• Wider testing because of large contact potential: 167 staff and pupils attending were tested. | 100/30 | Yes | 6 | 10/72 | 2 | 23 | 3/78 | 15 | Primary school outbreak involving staff and pupils, originally linked to a household cluster; evidence of wider transmission across bubbles. |

### Single, confirmed cases

The 67 single cases included 30 (45%) students and 37 (55%) staff and occurred mainly in primary schools (45/67, 67%) and early years settings (10/67, 15%) (**Figure 4a**). None of the children and one staff member was hospitalised with COVID-19 and required intensive care admission for respiratory support. One secondary school teacher died of COVID-19 after acquiring the infection from a household member with confirmed infection who had acquired the infection in the community. The teacher had not attended the school during their infectious period and, therefore, did not have contact with other staff or students within the school. Among 43/48 (90%) cases with available information, the case and contact bubble were excluded from 39 educational settings while four educational settings decided to close entirely because of a perceived risk of onward transmission, although this was contrary to national recommendations. For the remaining five cases (3 staff, 2 children) only the confirmed cases were isolated because they had remained outside the educational setting throughout their infectious period.

**Figure 4a.**
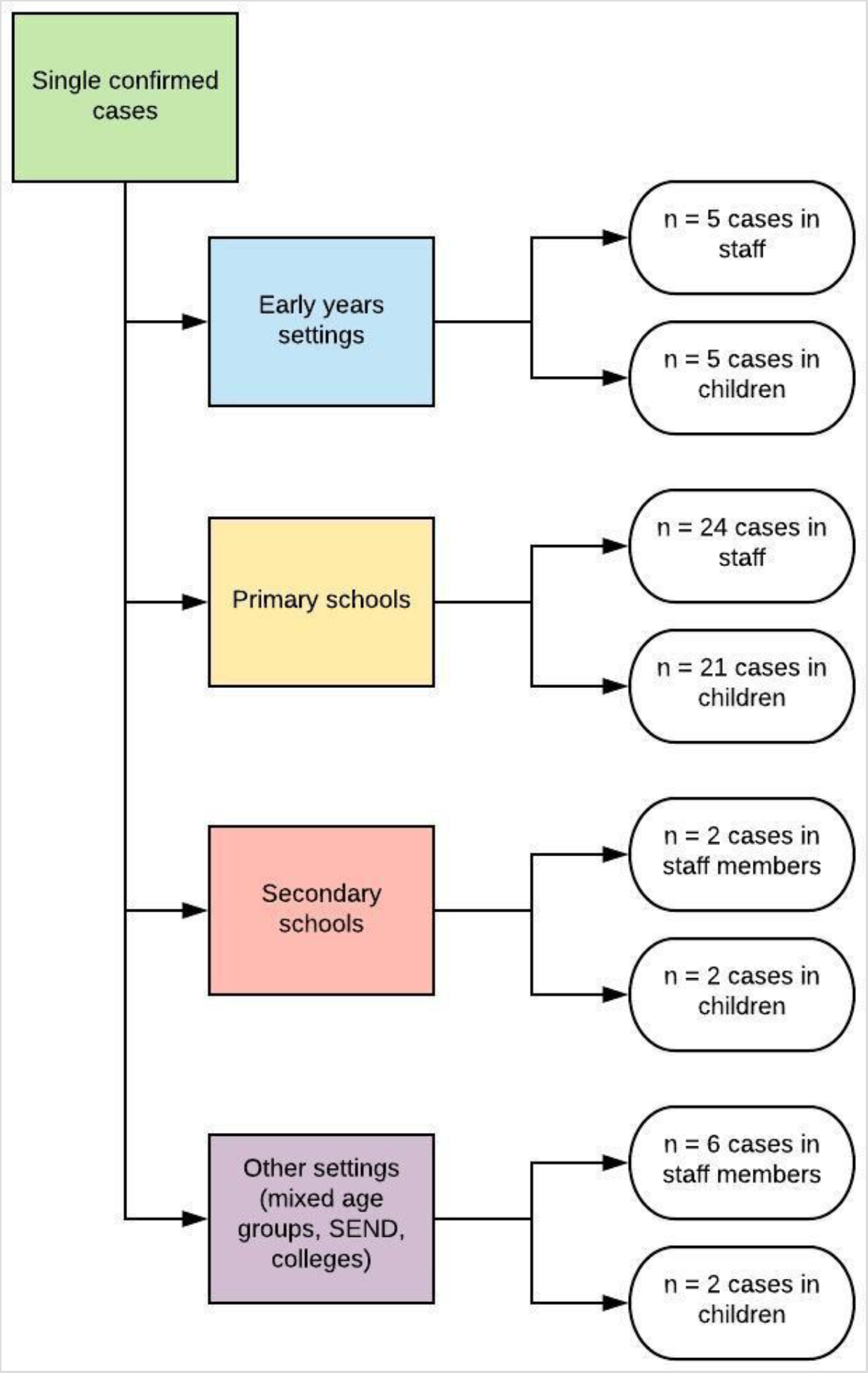
Summary of single confirmed cases of SARS-CoV-2 infection in staff and students by the type of educational setting in which they were reported

### Co-primary cases

There were 4 co-primary cases involving 10 children (**Figure 4b**). The co-primary cases lived in the same household, where they most likely co-acquired the infection. Three involved siblings in different primary schools (3, 2 and 2 siblings) and the fourth involved two separate household clusters (2 siblings and 1 child) in a preschool, with no contact links identified between the two households inside or outside the educational setting. All 10 children in the 5 households were asymptomatic and had been tested because they were household contacts of an index case (a parent in 4/5 households), who tested positive for SARS-CoV-2 after becoming symptomatic. Public health management included exclusion of the contact bubbles. In line with national recommendations, the settings remained open for the rest of the term and no additional cases were identified.

**Figure 4b.**
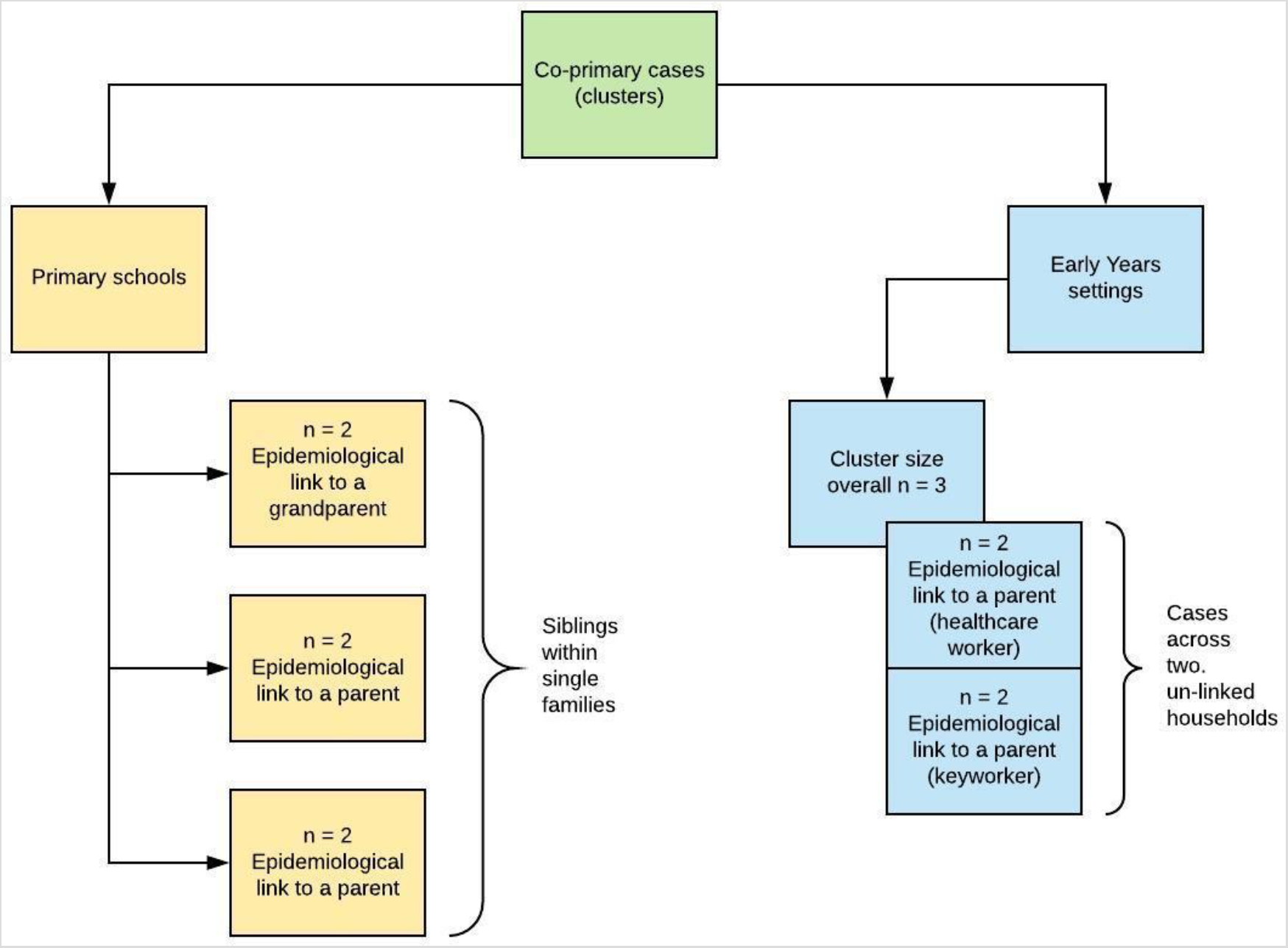
Summary of clusters involving co-primary cases by the type of educational setting in which they were reported. All co-primary cases were identified in students

### Confirmed outbreaks

There were 30 (18%) outbreaks with ≥2 confirmed cases in staff/pupils in primary schools (18, 60%), early years (7, 23%), secondary schools (2, 7%) and SEND schools (3, 10%) (**Figure 4c**). Affected contact bubbles were excluded from school in all 30 outbreaks and 13 (3%) also decided to close either on an interim basis (to allow for deep cleaning or for exclusion periods to elapse) or for the rest of the term.

**Figure 4c.**
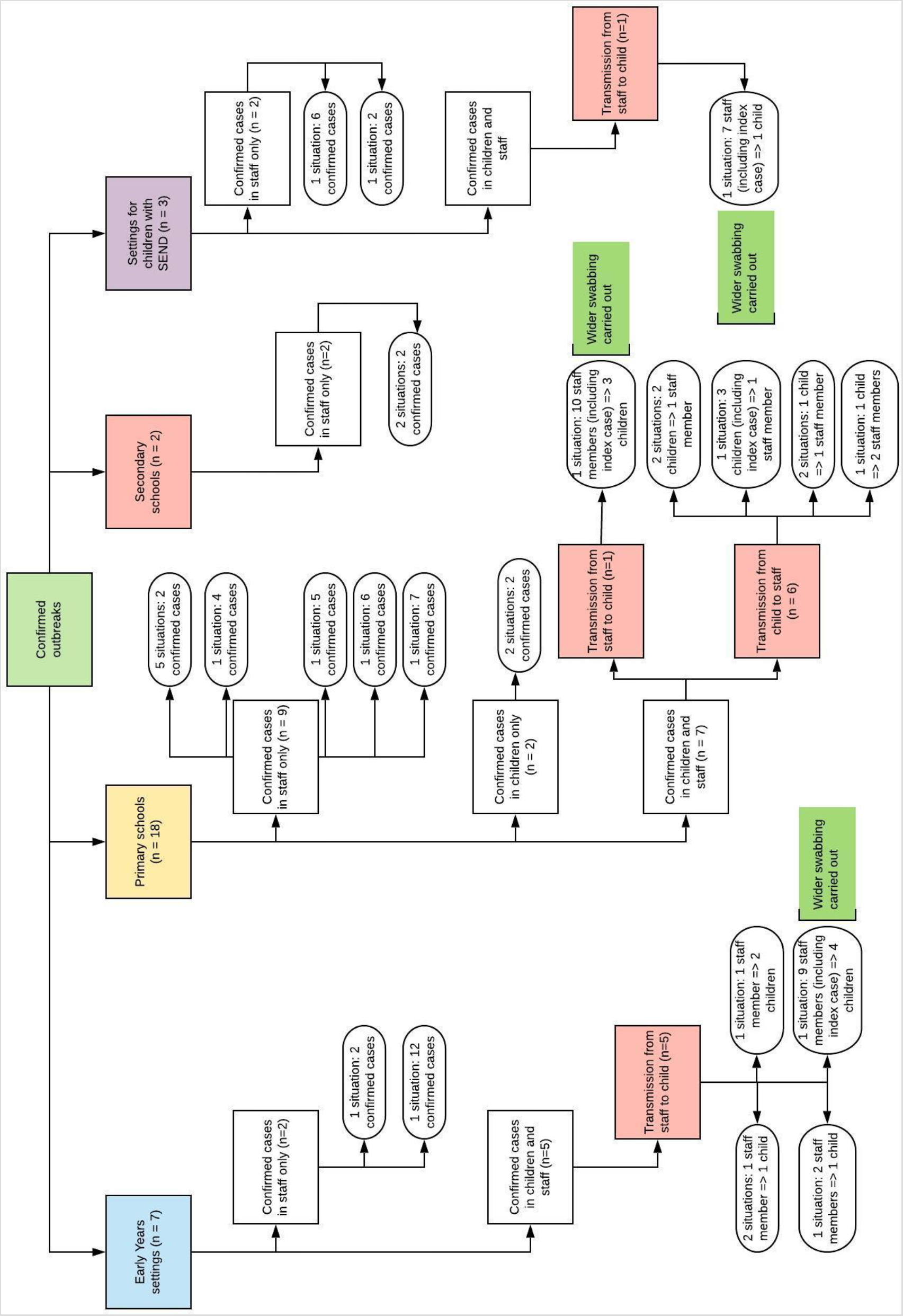
Summary of outbreaks by the type of educational setting in which they were reported, and likely direction of transmission between staff and students attending educational settings

Of the 18 primary school outbreaks, 9 involved staff only (affecting 32 members), including 5 where only two staff members were affected. In three of the remaining 4 staff outbreaks, the source was not identified but the outbreak was propagated through contact between administrative and teaching staff in school. The final staff outbreak was unusual in that 3 of the 4 SARS-CoV-2 positive staff members likely acquired their infections from household members with confirmed COVID-19.

Seven primary school outbreaks involved staff and students. In this small sample of outbreaks the student was most likely the index case in 6 outbreaks. In 5 outbreaks, the child (index case) was identified through ‘test and trace’; i.e. testing of the whole household when parents (healthcare workers in 3 cases) had tested positive for SARS-CoV-2. Seven staff members in contact with the index cases (3 tested because they became symptomatic) subsequently tested positive for SARS-CoV-2. A seventh outbreak involved potential staff-to-pupil transmission. A married couple who were staff members at the same school tested positive for SARS-CoV-2 after becoming symptomatic. Mass testing was undertaken because of the large number of potential school contacts and 8/74 (11%) additional staff and 3/93 (3%) children tested positive for SARS-CoV-2.

The final two primary school outbreaks involved possible transmission between two primary school children only, although no clear contact between the affected children within the educational setting could be established and acquisition in the household or community cannot be ruled out.

A staff member was identified as the index case in all 7 outbreaks in early years settings. Two outbreaks involved only staff members, including one where 12/16 staff tested positive for SARS-CoV-2 following community exposures linked to a religious festival. Wider swabbing in two outbreaks identified 9/43 (21%) children, 16/28 (57%) staff and 10/112 (9%) household contacts of staff/students as SARS-CoV-2 positive.

The remaining 5 outbreaks all involved staff members as the most likely index cases, with 19 confirmed cases among staff members and one in a child attending a SEND school, who potentially acquired the infection from a staff member with confirmed COVID-19. Wider swabbing in the SEND school was performed because of the perceived vulnerability of students and identified 6 additional SARS-CoV-2 positive cases in staff members (6/18, 33%) and one in a child (1/12, 8%). All five early ears settings excluded contacts of confirmed cases and two closed for deep cleaning and/or the rest of the term.

### Source of infection

The source of infection was not systematically collected for single confirmed cases. The 4 co-primary events involved 10 children who acquired the infection from a household member. The probable transmission direction for the 30 confirmed outbreaks was: staff-to-staff (n=15), staff-to-student (n=7), student-to-staff (n=6) and student-to-student (n=2). Of the 30 student cases involved in an outbreak, the potential source of infection in 27 children included a household contact (n=8), a school staff member (n=17) and another student (n=2). Among 91 staff members involved in an outbreak, where a potential source of infection could be identified, 9 acquired the infection from a household contact and were the likely index case in the outbreak and 52 likely acquired the infection in the educational setting from another staff member (n=46) or another child (n=6).

## DISCUSSION

Active, prospective and systematic national surveillance identified a low risk of SARS-CoV-2 infections or outbreaks among staff and students in educational settings during the first month of schools re-opening after easing of COVID-19 lockdown in England. Primary schools had the highest SARS-CoV-2 infection and outbreak rates among educational settings, likely reflecting the larger volume of children and staff returning to these educational settings during the summer mini-term. SARS-CoV-2 infection and outbreak rates were higher in staff than students and, among students, primary school children were more likely to be affected. Outbreaks involving staff-to-staff transmission were most common, whilst transmission between staff and students were less common and student-to-student transmission rare. Most school children with SARS-CoV-2 infection were identified as part of contact tracing after their parent, often a healthcare worker, was diagnosed with COVID-19. We found a strong correlation between community SARS-CoV-2 incidence and COVID-19 outbreaks in educational settings.

### Risk of infections and outbreaks

England took a cautious approach for opening schools as part of the easing of the lockdown, when community SARS-CoV-2 incidence was low across most regions specially when compared to the peak of the pandemic in mid-April 2020.^5^ Extensive social distancing and infection control measures were implemented for returning staff and students with strict limitations on the number of staff and students in each bubble. In addition, educational settings in regions with high incidence areas remained closed and parents were not required to send their children to school. The low rates of SARs-CoV-19 infection and outbreaks in England are consistent with the experience of other countries that opened their educational settings after lockdown as well as those that kept their preschools and primary schools open throughout the pandemic period, albeit with varying background incidence rates.^12^ There are very few reports of widespread infection or transmission of SARS-CoV-2 in preschools and primary schools, especially when compared to other institutions such as hospitals, care homes,^13,14^ prisons,^15^ and certain workplace settings.^16^ We did, however, find a strong correlation between community SARS-CoV-2 incidence and risk of outbreaks in educational settings, even during a period of low SARS-CoV-2 incidence. This is not surprising since increased community transmission provides more opportunities for virus introduction into educational settings.

The potential for spread within educational settings, as observed from the wider swabbing of some schools in our surveillance and from recent reports from other countries,^17^ does suggest that school closures may be necessary as part of lockdown in regions with increasing community infection, although given what is known about the detrimental effects of lack of access to education on child development, these should probably be considered only *in extremis* by comparison with other lockdown measures.^18^ Reassuringly, our findings indicated that early detection and isolation of staff and students can prevent progression to an outbreak in most cases, highlighting the importance of the ‘test, track, and trace’ approach.

### Risk of SARS-CoV-2

Within the educational setting, the higher risk of SARS-CoV-2 among staff highlights a need to strengthen infection control measures at two levels. Staff members need to be more vigilant for exposure outside the school setting to protect themselves, their families and the educational setting. Within the education premises, stringent infection control measures between staff need to be reinforced, including use of common staff rooms and cross-covering staff across bubbles.

Students, on the other hand, mostly acquired SARS-CoV-2 infection at home, usually from a keyworker or healthcare worker parent. Most children were asymptomatic and only identified as part of contact tracing after their parent developed COVID-19, highlighting the importance of access to rapid testing, reporting and contact tracing for individuals to protect the wider community. Reassuringly, we found very little transmission between the students which is consistent with emerging literature for preschool and primary school students,^19^ and, even among the coprimary cases, the siblings did not seed the infection into different bubbles. Additionally, there were very few transmission events between staff and students, especially given the difficulties in maintaining social distancing with younger children and those attending SEND schools.

### Strengths and weaknesses of the study

The main strength of this study is the public health infrastructure that allows real-time reporting, recording, risk assessment and management of situations across England. The large number of reports of suspected situations highlight the strong historical engagement between educational settings and HPTs. Daily assessment of situations of interest allowed a pragmatic approach to risk assessment, the need for additional investigations and infection control measures.

There are, however, important limitations when considering the generalisability of our findings. Educational settings opened after national lockdown when SARS-CoV-2 incidence was low and only in regions with low community transmission. Settings that opened had stringent social distancing and infection control measures in please and, in addition to school attendance not being mandatory, there were strict protocols for class and bubble sizes, which may not be achievable when schools opening fully in the next academic year (and indeed, updated schools guidance now recognises that bubble size may need to be increased from September to ensure that a full range of activities is feasible). Only 1.6 million of the 8.9 million students nationally attended any educational setting during the summer mini-term.^20^ Additionally, very few secondary schools opened (and those that did, did so with small class sizes) during the summer mini-term and our results, therefore, are not likely to be generalisable to secondary schools, especially since the risk of infection, disease and transmission is likely to be higher in older than younger children.^19,21^ Moreover, each situation was risk-assessed on a case-by-case basis and only a few settings were selected for wider testing. The risk-based approach was considered more pragmatic and allowed objective evaluation of the recommended national approach to isolate affected bubbles by monitoring the educational setting for additional cases.^7^ Identification of additional cases following wider testing highlights the importance of enhanced surveillance to better understand infection and transmission in educational settings. Finally, whole genome sequencing could help determine whether the outbreaks were caused by the same strain or due to multiple separate introductions into the educational setting and will be pursued for a sample of outbreaks where mass testing was undertaken.

### Conclusions

In conclusion, re-opening of schools was associated with very few outbreaks after easing of national lockdown in England. SARS-CoV-2 infection and outbreaks were more likely to involve staff members, highlighting a need to improve education and infection control measures for this group. The strong correlation between COVID-19 outbreaks and regional SARS-CoV-2 incidence highlights the importance of controlling the disease in the community to protect the staff and students in educational settings.

## Data Availability

Applications for relevant anonymised data should be submitted to the Public Health England Office for Data Release: https://www.gov.uk/government/publications/accessing-public-health-england-data/about-the-phe-odr-and-accessing-data

## Research In Context

### Evidence Before this study

We searched PubMed with the terms “COVID-19” or “SARS-CoV-2” with “school”, “education”, “nursery” or “student” to identify publications relating to COVID-19 cases and outbreaks in educational settings since January 2020. The majority of publications were reviews and opinion pieces on the impact of school closures on disease transmission and child health. There are very few reports of COVID-19 outbreaks in educational settings, mainly involving a single school with a limited number of staff and students affected. Secondary schools appear to experience wider transmission and larger outbreaks than schools for younger students.

### Added Value of This Study

In England, the reopening of mainly preschools and primary schools in June 2020 following the easing of national lockdown was associated with a total of 198 confirmed COVID-19 cases (70 in children, 128 in staff members), of which 67 were single confirmed cases (30 in children, 37 in staff) and 121 were linked to outbreaks (30 in children and 91 in staff). There were 30 outbreaks nationally, with an estimated 0.5, 4.8 and 1.6 outbreaks per 1,000 settings per month in early years, primary schools and secondary schools. The number of outbreaks in educational settings correlated strongly with regional COVID-19 incidence (0.51 outbreaks for each new SARS-CoV-2 infection per 100,000 in the community; p=0.001). Staff members were more likely to be affected than students.

### Implications of all the Available Evidence

The re-opening of schools was associated with very few COVID-19 outbreaks after easing of national lockdown in England. SARS-CoV-2 infection and outbreaks were more likely to involve staff members, emphasising a need to improve education and infection control measures for this group both within and outside the educational setting. The strong correlation between COVID-19 outbreaks and regional SARS-CoV-2 incidence highlights the importance of controlling the disease in the community to protect the staff and students in educational settings.

## Acknowledgements

We are grateful to Jemma Walker, Senior Statistician with the National Infection Service at PHE for assistance with statistical analyses in the paper.

## Ethical approval

PHE has legal permission, provided by Regulation 3 of The Health Service (Control of Patient Information) Regulations 2002, to process patient confidential information for national surveillance of communicable diseases and as such, individual patient consent is not required.

## REFERENCES

1. Munro APS, Faust SN. Children are not COVID-19 super spreaders: time to go back to school. Arch Dis Child 2020.

2. Viner RM, Russell SJ, Croker H, et al. School closure and management practices during coronavirus outbreaks including COVID-19: a rapid systematic review. Lancet Child Adolesc Health 2020;4:397–404.

3. Viner RM, Bonell C, Drake L, et al. Reopening schools during the COVID-19 pandemic: governments must balance the uncertainty and risks of reopening schools against the clear harms associated with prolonged closure. Arch Dis Child 2020.

4. Viner RMM, O.T.; Bonell, C.; Melendez-Torres, G.J.; Ward, J.L.; Hudson, L.; Waddington, C.; Thomas, J.; Russell, S.; van der Klis, F.; Panovska-Griffiths, J.; Davies N.G.; Booy, R.; Eggo, R. Susceptibility to and transmission of COVID-19 amongst children and adolescents compared with adults: a systematic review and meta-analysis. MedRxiv Posted 24 May 2020.

5. National COVID-19 surveillance reports 2020. at https://www.gov.uk/government/publications/national-covid-19-surveillance-reports.)

6. Prime Minister’s statement on coronavirus (COVID-19): 23 March 2020. 23 March 2020. (Accessed 05 August 2020, 2020, at https://www.gov.uk/government/speeches/pm-address-to-the-nation-on-coronavirus-23-march-2020.)

7. Guidance for full opening: schools. 27 July 2020. (Accessed 05 August 2020, at https://www.gov.uk/government/publications/actions-for-schools-during-the-coronavirus-outbreak/guidance-for-full-opening-schools)

8. Children with special educational needs and disabilities (SEND). 2020. (Accessed 05 August 2020, at https://www.gov.uk/children-with-special-educational-needs.)

9. COVID-19 testing data: methodology note (updated: 29 July 2020). 2020. (Accessed 05 August 2020, 2020, at https://www.gov.uk/government/publications/coronavirus-covid-19-testing-data-methodology/covid-19-testing-data-methodology-note.)

10. The national curriculum. 2020. (Accessed 05 August 2020, at https://www.gov.uk/national-curriculum.)

11. Attendance in education and early years settings during the coronavirus (COVID-19) outbreak: 23 March to 9 July 2020. 2020. (Accessed 05 August 2020, 2020, at https://www.gov.uk/government/statistics/attendance-in-education-and-early-years-settings-during-the-coronavirus-covid-19-outbreak-23-march-to-9-july-2020.)

12. Levinson M, Cevik M, Lipsitch M. Reopening Primary Schools during the Pandemic. N Engl J Med 2020.

13. Graham N, Junghans C, Downes R, et al. SARS-CoV-2 infection, clinical features and outcome of COVID-19 in United Kingdom nursing homes. J Infect 2020.

14. Arons MM, Hatfield KM, Reddy SC, et al. Presymptomatic SARS-CoV-2 Infections and Transmission in a Skilled Nursing Facility. N Engl J Med 2020.

15. Franco-Paredes C, Jankousky K, Schultz J, et al. COVID-19 in jails and prisons: A neglected infection in a marginalized population. PLoS Negl Trop Dis 2020;14:e0008409.

16. Waltenburg MA, Victoroff T, Rose CE, et al. Update: COVID-19 Among Workers in Meat and Poultry Processing Facilities – United States, April-May 2020. MMWR Morb Mortal Wkly Rep 2020;69:887–92.

17. Stein-Zamir C, Abramson N, Shoob H, et al. A large COVID-19 outbreak in a high school 10 days after schools’ reopening, Israel, May 2020. Euro Surveill 2020;25.

18. Auger KA, Shah SS, Richardson T, et al. Association Between Statewide School Closure and COVID-19 Incidence and Mortality in the US. JAMA 2020.

19. Park YJ, Choe YJ, Park O, et al. Contact Tracing during Coronavirus Disease Outbreak, South Korea, 2020. Emerging Infectious Disease journal 2020;26.

20. Academic Year 2019/20: Schools, pupils and their characteristics. 22 July 2020. (Accessed 05 August 2020, at https://explore-education-statistics.service.gov.uk/find-statistics/school-pupils-and-their-characteristics.)

21. Gudbjartsson DF, Helgason A, Jonsson H, et al. Spread of SARS-CoV-2 in the Icelandic Population. N Engl J Med 2020.

